# A scoping review on the associations and potential pathways between malnutrition and measles

**DOI:** 10.1101/2023.01.21.23284872

**Authors:** Isabelle CT Tran, Christopher Gregory, Patrick O’Connor, Annette Imohe, Lien Anh Ha Do, Parminder S Suchdev

## Abstract

Amid a measles resurgence worldwide, many affected regions also find themselves in circumstances of conflict, humanitarian or political crises, climate extremes, ongoing pandemic challenges, income inequality and economic downturns. Those contextual factors have driven increases in worldwide undernourishment over the past year. The overlap and frequent occurrence of those socio-structural determinants in fragile geographies is worrying as a synergistic relationship between measles and malnutrition has been reported. A scoping review was conducted to map current evidence and identify research gaps on the association between malnutrition and measles.

Sixty-seven studies were retained following a search of multiple databases, snowballing and consultations with experts. Studies reported on a measure of association, a relationship, or potential mechanisms behind the link between malnutrition and measles.

Overall, there was evidence of a positive correlation between undernutrition and reported measles incidence and mortality. All studies reviewed found an association between vitamin A deficiency and increased likelihood of incident measles, measles-related complications and measles-related deaths. Available evidence also suggested that therapeutic vitamin A can significantly reduce the odds of measles-related mortality, and preventative vitamin A can reduce reported measles incidence. Little data shed light on potential pathways behind malnutrition and measles. Inconsistent variable definitions (i.e. malnutrition and measles cases) across studies precluded calculating a cumulative effect size, and findings may be impacted by uncontrolled confounding factors.

This scoping review reinforces the hypothesis of a synergistic relationship between measles and malnutrition and highlights the need for an integrated approach to the delivery of malnutrition interventions and measles prevention and control. In addition, further robust studies are needed to better define pathophysiological targets for interventions and allow for stronger inferences to policy making. Future research should also consider using a standardized approach to defining malnutrition and measles to ensure comparability of outcomes across studies.

## Introduction

Measles is one of the most contagious respiratory infections, spread from person-to-person via airborne transmission.(1) Malnourished children with weaker immune systems are particularly at risk of developing debilitating complications.(1) Following the introduction of widespread measles immunization campaigns, cases dropped 95% over a 35-year period, from 1980 to 2015. (1) From 2000 to 2016, measles deaths dropped by 84%. (1) Despite this progress, global vaccination rates have been falling short of universal coverage targets. For example, while 95% coverage for two doses of measles immunization is required to achieve disease elimination, coverage has stagnated at around 10% below the recommended threshold for the first dose, and 25% for the second. (2, 3). From 2016 to 2019, there was a measles resurgence worldwide: incident cases rose by 556%. (3) In that same period of time, measles-related deaths surged nearly 50%. (3) In the recent years, past achievements have been lost largely due to a failure to vaccinate, driven by limited access to health services and/or vaccine hesitancy. (3) The 2020 global COVID-19 pandemic also had repercussions on immunization services. The latest World Health Organization (“WHO”) and United Nations International Children’s Emergency Fund (“UNICEF”)’s estimates found that global coverage of the first dose for measles dropped 2%, from 86% in 2019 to 84% in 2020, and coverage of the second dose for measles dropped 1%, from 71% in 2019 to 70% in 2020. These decreases have led to an additional 3 million potentially unvaccinated children in 2020 compared to 2019. (4)

The largest and most disruptive outbreaks in 2020-21 were across the African region, as well as in Afghanistan, Yemen and Pakistan. (5) Many of the affected countries are currently in a context of conflict or humanitarian emergency, with weakened public infrastructures, disruptions to immunization delivery and limited access to healthcare. Several of them are also being affected by climate extremes, income inequality and economic downturns. All those contextual drivers have led to an increase in worldwide undernourishment over the past year. In 2020, Africa and Asia were home to more than 90% of the children with wasting and/or stunting. (6) UNICEF analyses estimate that, from 2020 to 2021, coverage of essential nutrition services, such as early detection and treatment of wasting, dropped by at least 10-24% in a fifth of countries which reported data. From July 2020 to July 2021, national coverage of vitamin A supplementation, a World Health Organization recommended intervention that is critical to maintain strong immune systems and improve a child’s survivability, dropped in 24 countries.(7) Though the impact of the pandemic on global malnutrition has not yet been fully assessed, initial projections suggest that the prevalence of moderate or severe wasting among children under 5 years old could increase by 14.3% due to COVID-19 repercussions.(8) This is equivalent to an additional 6.7 million wasted children, with approximately 57.6% of them in Asia and 21.8% in Africa.(8) Current Russo-Ukrainian conflicts also have impacts on the global food supplies, further exacerbating global food insecurity and malnutrition.(9)

The coincidence of measles and undernutrition in fragile contexts is worrying as undernourished children are at higher risk of measles-related complications and deaths. (1) Measles has also often been reported as a potential compounding factor for malnutrition. The infection can cause severe diarrhea, contributing to a loss of nutrients and body fluids. (1) In addition, measles is commonly complicated by mouth ulcers and gastroenteritis. (10) This can cause discomfort with eating, decreased appetite and reduced dietary intake, and all of these changes are occurring on a backdrop of increased energy and nutrient requirements due to physiological changes caused by the illness. Potential complications also include erasure of pre-existing immune memory cells and immune suppression, which can last several weeks and up to 2-3 years following an acute measles episode.(11) Infected individuals are therefore more susceptible to secondary infections, compounding the risk of nutritional deficiency.(11) Finally, noma, a debilitating condition characterized by gangrenous ulcerations of the mouth and face, has been associated with severe measles. (12) Noma cases have been described particularly among malnourished, young children living in extreme poverty and in countries with low measles vaccination rates. (12)

However, little is known about the causal mechanisms behind the potential association between measles and malnutrition. While malnutrition in all its forms may include undernutrition, hidden hunger (i.e. deficiencies of essential vitamins and minerals), and overweight/obesity, for the purpose of this review we focused on undernutrition and micronutrient deficiencies.(13) This paper aims to explore the current state of knowledge regarding the association between undernutrition and measles, and to highlight any evidence gaps.

## Review questions

- Does malnutrition impact morbidity, mortality, and incidence of measles, and to what extent?
- Does measles cause or contribute to malnutrition, and to what extent?
- What are the potential pathways underlying the relationship between measles and malnutrition?

## Methodology

### Selection of articles

An exploratory review was conducted from July 2021 to February 2022, using various combinations of the following search terms in all fields: “measles”, “malnutrition”, “malnourished”, maln*”, “undernutrition”, and “undernourish*”. This search query was conducted in Pubmed and Google Scholar’s databases and yielded 822 articles. Following feedback from senior research supervisors, the search was updated in March 2022 with keywords relating to “vitamin A”, “retinol”, “serum retinol”, “hyporetinemia” and measles. This update was limited to entries from 2018 to 2022 and yielded 55 additional articles. A sample search strategy for Pubmed can be found in S1 File.

After the identification process, the remaining articles’ abstracts were screened and, where an abstract was unavailable, full texts scanned through, to determine eligibility. As this review focuses on the association between malnutrition and measles, studies that examined another primary variable were excluded (e.g., diseases other than measles). Noma, in particular, was the main topic in 196 articles, and those studies were excluded during screening.

To be included in the review, a study had to

- Report on a measure of association, a relationship or potential mechanisms behind the link between malnutrition and measles; and
- Be published in English or French.

Case series, editorials, commentaries, animal studies and studies that could not isolate the effect of measles or malnutrition, were excluded (i.e., effect size assigned to multiple, concurrent causes).

Five additional studies were identified by snowballing and through consultations with experts.

Following the Preferred Reporting Items for Systematic Reviews and Meta-Analyses (“PRISMA”) flow diagram’s criteria presented in Fig 1, 67 studies were included in the review.

**Fig 1.**
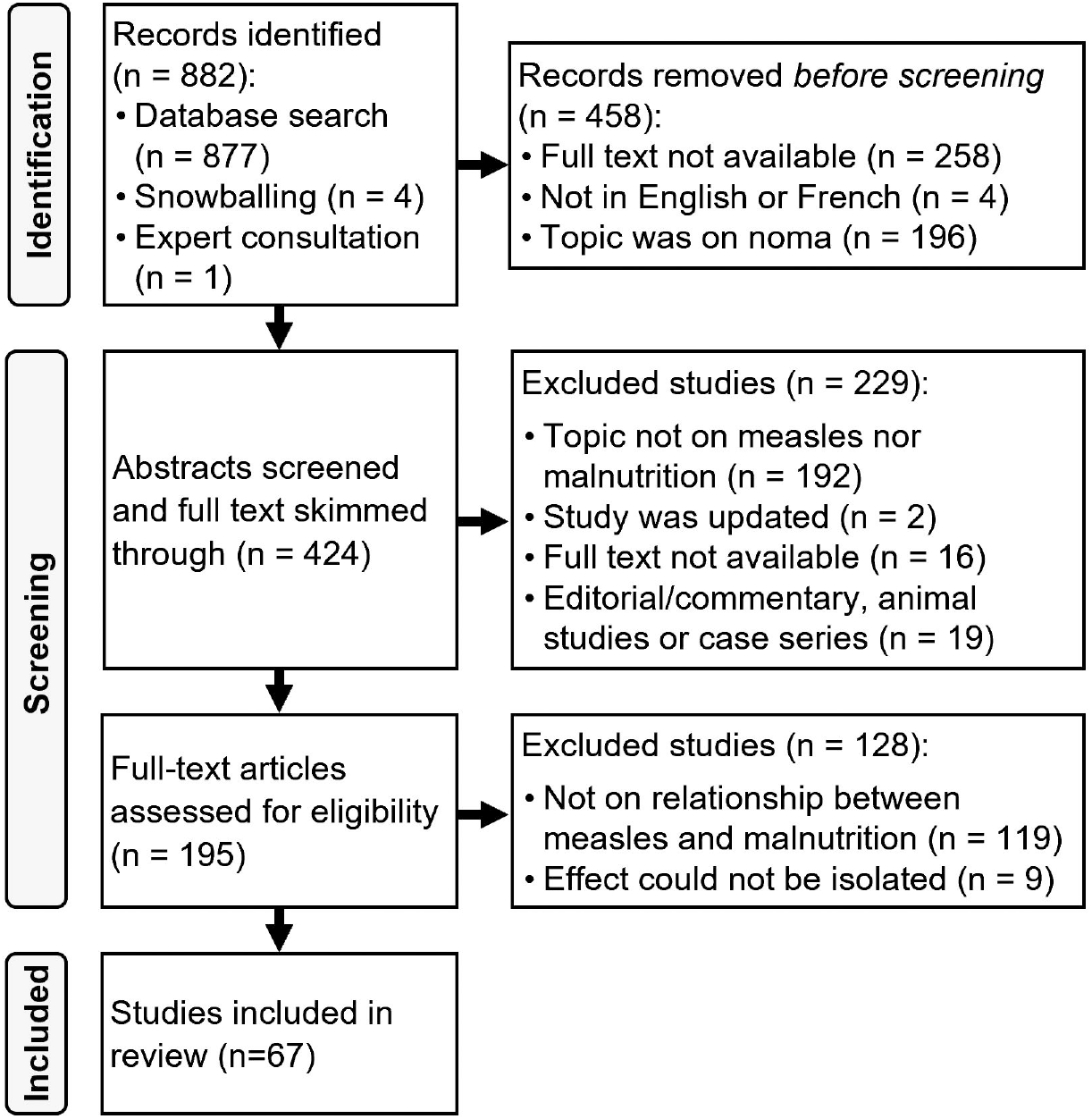
PRISMA Flow Diagram of the Study Selection Process. Snowballing refers to using the references or citations of a record to identify further studies of interest.

This review followed the reporting guidelines of the Preferred Reporting Items for Systematic Reviews and Meta-Analyses extension for Scoping Reviews (“PRISMA-ScR”) checklist (see S2 File). As it aimed to explore the key characteristics around the malnutrition-measles relationship and to identify current evidence gaps, the scoping review approach was selected over the systematic review.(14)

### Expert consultations

Expert consultations were held during August 2021 to identify any potentially missing relevant literature and to obtain critical input on important areas of discussion.

Four experts in the fields of immunology, respiratory and tropical medicine, respiratory viruses, nutrient metabolism and tropical pediatrics, and global health and micronutrient malnutrition, were invited. Experts were identified through a snowballing approach, expanding from the professional networks of two senior research supervisors.

In March 2022, a final draft manuscript was sent out to these experts for feedback.

### Study characteristics

Studies that fulfilled the inclusion criteria are presented in S1 Table. Much of the evidence on the relationship between measles and malnutrition is based on older studies, whereas studies from 1991 onwards tend to look at vitamin A supplementation and deficiency.

The majority of the articles retained were published more than 10 years prior (See Fig 2):

**Fig 2.**
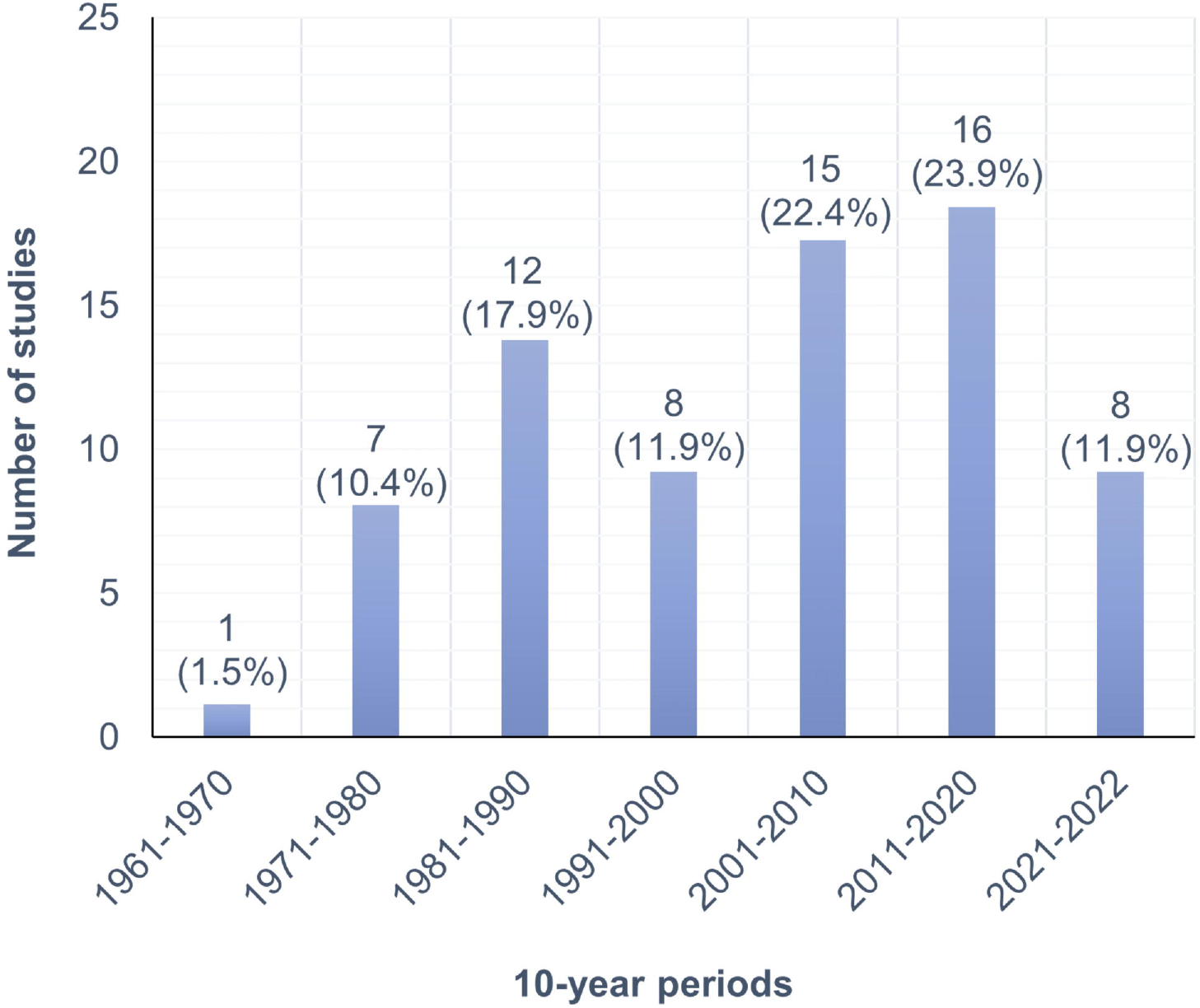
Number of studies published per 10-year period about measles and malnutrition.

22 studies were reviews, of which 6 included a meta-analysis, and 45 were primary research reports with the following study designs:

- Descriptive studies (10);
- Analytical observational studies (i.e., cohort and case-control studies) (32); and
- Analytical experimental studies (i.e., randomized controlled trials (“RCT”)). (3)

Data from retained studies was extracted by using a charting table in Excel. This process was iterative, and the variables to extract were continuously updated following feedback from expert consultations and senior reviewers. Data was charted for the following variables: author(s), year of publication, type of study, setting, epidemiologic setting, independent variable(s) of interest, dependent variable, aspect of measles studied, measure of association, determination of a measles case, indicator of vitamin A deficiency (if applicable), vitamin A supplementation regimen (if applicable), malnutrition nomenclature, definition of a malnourished case.

Articles were grouped according to the main variables (i.e. measles, malnutrition, vitamin A deficiency, vitamin A supplementation) they studied. All charted data was synthesized in supplementary tables (see S1 Table to S5 Table).

### Definitions of malnutrition

The term “malnutrition” involves a large spectrum of nutritional impairments, including deficiencies, and excesses or imbalances in energy and/or nutrient intake. As the publication period of included studies ranges from 1969 to 2021, definitions of a malnourished case varied widely. Some of the nomenclature is no longer in use. S2 Table presents the distribution of different malnutrition (undernutrition) definitions across the studies reviewed.

To ensure that the evidence from this review can be translated into practice, findings are categorized according to the most current definitions of malnutrition, as per UNICEF’s classification (13):

- Undernutrition (i.e., macronutrient deficiencies);
- Micronutrient deficiencies; and
- Overnutrition

As overnutrition was not investigated in the retained studies, this paper will refer to malnutrition in the sense of undernutrition and micronutrient deficiencies. In terms of micronutrient status, all articles examined vitamin A deficiency and supplementation, save for two studies which looked at zinc supplementation.

The earliest label for malnutrition was “protein-calorie malnutrition (PCM)”, which came into use in the 1960s. (15) Half of the articles that defined malnutrition as PCM, assessed the condition via weight-for-age standard deviations. The remaining articles did not specify how PCM was assessed.

In the 1970s, “protein-calorie malnutrition” transitioned into “protein-energy malnutrition (PEM)”. (16) Under the WHO and the Food and Agriculture Organization (“FAO”) definitions, PEM encompassed “a range of pathological conditions arising from coincident lack, in varying proportions, of protein and calories, occurring most frequently in infants and young children, and commonly associated with infections”. (16) In the literature, PEM generally refers to two main syndromes defined by specific clinical signs and symptoms: kwashiorkor and marasmus. Kwashiorkor refers to a form of oedematous malnutrition, characterized by skin depigmentation and sloughing, and fatty liver. (17) Marasmus refers to a loss of subcutaneous fat and muscle atrophy and is often seen as a wasting syndrome. (17) “Marasmic kwashiorkor”, representing clinical features from both marasmus and kwashiorkor, was examined in one of the studies reviewed. In total, 9 studies used some variation of the PEM nomenclature.

Wasting and stunting are defined by the WHO as “weight for height as an indicator of the present state of nutrition (i.e. acute malnutrition / wasting) and height for age as an indicator of past nutrition (i.e. chronic malnutrition / stunting)”. (18). Some studies also measured malnutrition as being “underweight”, a composite of stunting and/or wasting.

Most studies defined stunting, wasting and underweight as per the WHO’s anthropometric standards. (19) Stunting was defined as a height-for-age Z-score (HAZ) below − 2 (i.e. more than 2 S.D. below the population median), wasting was defined as a weight-for-height Z-score below −2, and underweight was defined as weight-for-age Z-score below −2. (19) In three historical studies from the 1970s, however, Harvard standards were used instead and expressed degrees of malnutrition according to percentages below a standard median (i.e. 90%, 80%, 70%, 60%). (20–22) Six studies also used mid-upper arm circumference (MUAC) as an indicator for severe acute malnutrition (“SAM”) in infants and children 0 to 95 months. (23–28)

### Definitions of a measles case

A measles case is typically identified through clinical signs and symptoms, laboratory confirmation or epidemiological linkage.(29)

The WHO classifies measles cases in three categories:(29)

1. Suspected case: presence of fever and characteristic rash, or an individual suspected by a health-care worker to be infected with measles.
2. Clinically compatible measles: presence of fever, characteristic rash and either symptoms of coughing, coryza and/or conjunctivitis.
3. Confirmed case:

a. Epidemiologically linked: a suspected case of measles geographically and temporally linked with dates of rash onset occurring 7–23 days apart from a laboratory-confirmed or another epidemiologically linked measles case
b. Laboratory confirmed: IgM antibody detection for measles by enzyme immunoassay, detection of measles-specific RNA by RT-PCR or viral isolation by cell culture.

In this review, 29% (16/56) of the studies identified measles cases through the use of clinical signs and symptoms, 18% (10/56) used serological testing, of which 4% (2/56) also included epidemiological linkage, and 54% (30/56) did not specify how measles cases were defined.

In addition, clinical signs and symptoms used to define a case tended to vary in older historical studies. One study defined severe measles as the manifestation of unusually severe inflammatory change in a tissue known to be infected with measles (e.g.: extensive epidermal desquamation, severe keratitis, conjunctivitis) and two others identified cases through maternal and community reporting by asking about measles by its local name.

S3 Table presents the distribution of measles definitions across the studies reviewed.

### Definitions of vitamin A deficiency

Cases of vitamin A deficiency are primarily identified using biochemical indicators (i.e. serum retinol, breast milk retinol and/or serum retinol binding protein) and/or clinical eye signs (e.g. Bitot’s spots, corneal xerosis, keratomalacia, corneal scars).(30) Biomarkers are used as surrogate indicators of liver retinol reserves, which are considered as the gold standard to assess vitamin A status. Overt clinical damage to the eye are typically late manifestations of vitamin A deficiency.(30) However, several biomarkers of vitamin A status, such as retinol and retinol binding protein, may be confounded by the effects of inflammation, as they are themselves negative acute phase proteins.(31)

From 1963 to 1996, international guidance for serum retinol cut-off values recommended any concentrations below 10 µg/100 ml, or 0.35 µmol/L, among children younger than 6 years old, to be indicative of a deficiency.(32) In 1992, that cut-off was considered too low to identify asymptomatic individuals with subclinical vitamin A deficiency. (32) The recommended cut-off was modified to concentrations below 20 µg/100 ml, or 0.70 µmol/L, among children 6-71 months of age, in 1996. (32)

Six out of the fifteen studies on vitamin A deficiency in this review used serum retinol as indicator (See S4 Table). While three of them were published prior to 1996, the serum retinol threshold in those studies was 0.70 µmol/L, as per current guidelines. Two studies used clinical eye signs, one used conjunctival eye cytology, another used the molar ratio of retinol-binding protein to transthyretin, and 5 articles did not specify how vitamin A deficiency was assessed.

### Vitamin A supplementation approaches

This review considers two types of vitamin A supplementation approaches: preventative and therapeutic.

A typical preventative program includes providing two high-dose vitamin supplements each year among children 6-59 months, in areas with high under-5 mortality.(33)

Current measles management guidelines recommend providing therapeutic vitamin A supplementation to children under 5 years old who are infected with measles, as well as to adult measles patients with a suspected vitamin A deficiency. (34)

S5 Table presents the characteristics of all vitamin A supplementation studies included in this review.

## Findings on the association between measles and malnutrition

Studies investigated three different aspects of measles: incidence, mortality, and/or morbidity and its associations with malnutrition status.

### Vitamin A and susceptibility to measles infections

#### Vitamin A deficiency

In 1987, a community study in Ethiopia found that vitamin A-deficient children were 4.7 times more likely to develop measles (p = 0.01) than children that were not vitamin A-deficient. (35)

#### Preventative vitamin A supplementation

In 1997, a community based RCT found that the self-reported rate of measles cases was lower, although non-significantly (p=0.33), among children supplemented with vitamin A (100,000 international units (“IU”) for children aged 6-11 months or 200,000 IU for older children).(26) However, measles cases were identified through maternal reports, and the authors indicate that other diseases which cause rash and fever could have erroneously been reported as measles.(26)

A 2011 meta-analysis of 43 randomized controlled trials conducted between 1976 and 2010 found that vitamin A supplementation (i.e., varying dosage from 10 000 IU to 200 000 IU) was associated with a 50% reduced incidence of measles (95% confidence interval (“CI”): 0.37 to 0.67). (36) On the Grading of Recommendations, Assessment, Development and Evaluations (“GRADE”) scale, this evidence was considered as being of high quality. (36) This finding was reinforced by two Cochrane reviews in 2017 and 2022, which also concluded that vitamin A supplementation could reduce measles incidence by 50%, albeit their GRADE rating was reduced to moderate quality. (37, 38) This review included the same studies as the 2011 meta-analysis. The authors did not detail why the quality of evidence was modified; however, GRADE involves some subjective judgments and different sets of evaluators may rate a body of evidence differently.(39)

In 2021, a research report in India found that children 12 to 59 months old who received preventative vitamin A supplementation were 77% less at risk of getting measles (95% CI: 0.08-0.66).(40)

### Measles and increased susceptibility to vitamin A deficiency

Three studies found that measles can lead to or exacerbate vitamin A deficiency. (41–43) However, only one of these studies quantified the extent of measles’ influence: in India, children infected with measles were 9 times (95% CI: 2.75 – 31.19) more likely to develop subclinical vitamin A deficiency after the infection.(43)

### Bi-directional relationship between undernutrition and measles infections

Several community- and hospital-based studies reported significant associations between measles incidence and undernutrition. (23, 42, 44-48) In Yemen, suspected measles infections were three times higher among SAM cases (4.5%, p< 0.00) than in adequately nourished (1.4%) and in moderately malnourished (1.2%) children. (44) Another community study found that the identification of measles cases with fever and skin rash symptoms during an outbreak was 24.9 times more likely among malnourished children (i.e. aged 6 to 60 months) (95% CI: 1.9-329.6).(23) In Romania, reported measles cases were 3.4 times more likely than controls to have had a history of malnutrition (95% CI: 1.1-9.9). (45) In 2018, modelling from sub-national regions of Namibian aggregated measles incidence data from 2005 to 2014 found the risk of measles to be associated with the prevalence of underweight children under 5 years of age (95% CI: 0.0019, 0.1128).(49)

Additionally, measles was also found to cause weight loss in young children, more than other acute diseases. (47) Severely underweight children (weight-for age <− 3 standard deviations) were 1.7 times more likely to have had a history of measles in the preceding 6 months (95% CI: 1.3 – 2.4). (46) In Pakistan, measles cases < 9 months old were three times more likely to be malnourished than older children (95% CI: 1.2-7.5). (48)

### Malnutrition and measles-related mortality

In 2021, data from the Global Burden of Disease study database identified four main risk factors for measles-related deaths: wasting, underweight, stunting, and vitamin A deficiency.(50)

#### Vitamin A deficiency

A pooled analysis of 134 population-based surveys from 83 countries found that vitamin A-deficient people were 1.26 times more likely to die from measles (95% CI: 0.84–1.87), though this result was not statistically significant. (51) In those countries, 11, 200 measles deaths were estimated to be attributable to vitamin A deficiency in 2013. (51)

#### Therapeutic vitamin A supplementation

In 2005, a review of RCTs in which vitamin A therapy in varying doses was initiated after measles diagnosis found that vitamin A supplementation led to no significant reduction in the risk of mortality. (52) The authors highlighted, however, that factors such as greater vitamin A doses, hospitalization, age and geographic area, defined by the case-fatality rate in the region, could impact the effect size observed. (52) Within the same review, the subset of studies that found a statistically significant 64% reduction in the risk of mortality, all looked at vitamin A supplementation given in doses of 200,000 IU on two successive days, hospitalized patients, children under 2 years old, and were set in areas where the case-fatality rate was more than 10%. (52) Those factors are notable across previously published studies.

Trials from the 80s to the early 2000s in which a reduction in mortality is noted were generally hospital-based. (53–57) However, vitamin A in hospitalized patients does not consistently lead to a significant decrease in mortality risk, and effect sizes seem to vary depending on age, the type of complications and the degree of pre-existing malnutrition. (53, 56, 57) A 1992 review of hospital-based clinical trials found that two doses of 200,000 IU vitamin A given on successive days, or 20,000 IU daily for one to three weeks, reduced measles-related mortality in hospitalized children, however the effect was most significant in children under 2 years old. (56, 57) In India, patients with post-measles complications were 56% less likely to die when supplemented with 200 000 IU vitamin A on 2 successive days. (53) Those findings are consolidated in a 2002 meta-analysis which found that patients who received two doses of 200,000 IU vitamin A given on 2 consecutive days had 64% significantly lower overall mortality (Risk ratio (“RR”): 0.36; 95% CI: 0.14-0.82).(58) This impact was more important among children under 2 years old, where overall mortality risk dropped by 83% (RR: 0.17; 95% CI: 0.03-0.61).(58)

In 2 other meta-analyses of hospital-based studies, vitamin A supplementation significantly decreased the odds of mortality by 54% (95% CI: 18% - 72%) to 61% (95% CI: 22% – 66%). (54) (55) In both studies, vitamin A dosage was highly variable (see S4 Table).

In community-based articles, a 2021 study conducted during an outbreak in the Philippines found that children who did not receive vitamin A supplementation during measles episodes had significantly higher odds of mortality, nearly two-fold (Odds ratio (“OR”): 1.93; 95% CI: 1.31-2.85).(59) Compared to those who were treated with vitamin A, not receiving vitamin A supplementation increased the odds of death nearly two-fold (OR: 1.93; 95% CI: 1.31-2.85).(59) In 2017, another outbreak investigation found that children who did not receive vitamin A supplementation during measles illness had 2.3 times (95% CI: 0.74-7.4) increased risk, though non-significant, of mortality. (60) One 1997 study found no significant difference (p=0.51) in acute measles case fatality between children supplemented with 100 000 to 200 000 IU vitamin A (15.4%) and non-supplemented children (14.5%).(26)

### Preventative vitamin A supplementation

In a 1993 meta-analysis of community trials, the effect size was small but statistically significant in one study: children who received periodic vitamin A supplementation (i.e. 810 retinol equivalents per gram of dietary monosodium glutamate, a sodium salt used as flavor enhancer, or 8333 IU to 200 000 IU given once or every 4 to 6 months) were 25% to 30% less likely to die from measles. (55) Age was also an influential factor: vitamin A seemed more effective among younger children, though the specific age threshold was not specified. (55) In 1997, another review of 4 community studies indicated that vitamin A prophylaxis reduced measles mortality by about 50%. (61) The risk ratios in those studies, however, showed high variability (i.e. 0.58, 0.24, 0.67 and 0.82), where vitamin A would decrease the risk of measles mortality by 42%, 76%, 33% and 18%. (61)

In 2015, a pooled analysis of randomized trials found that vitamin A supplementation as a preventative intervention in community settings was associated with a 14% (95%: −8% - 32%)non-significant reduction in measles mortality. (51) Similarly, in 2017 and 2022, two Cochrane reviews concluded that vitamin A supplementation did not have a significant effect on mortality due to measles, however this evidence was graded as having low certainty. (37, 38)

#### Undernutrition

Out of 11 studies, 7 supported an association between undernutrition and measles mortality, (50, 62–67) 2 found no association, (68, 69) and two reviews found the evidence inconclusive.(70, 71)In 2004, a review of cohort studies found that 44.8% of measles-related deaths in young children were attributable to undernutrition. (64) In 2021, the most recent estimations of global measles burden evaluated that more than 50% of measles-related deaths were attributed to childhood wasting in high and middle-high socio-demographic index regions.(50) The socio-demographic index is an indicator of development status strongly correlated with health outcomes, and is a composite average of the incomes per capita, years of schooling and fertility rates in a country. (50) In addition, the study estimated that this proportion has been gradually increasing since 1990.(50) Among Somali refugees, acute malnutrition was an independent risk factor for measles mortality: malnourished children were 7.6x more likely to die from measles infections. (62) 3 other studies also noted that malnourished children had higher measles case-fatality rates, though only 1 of them quantified the extent to which it was higher (63, 65, 67) In Zimbabwe, stunted children had a case-fatality rate 1.7 times higher than children with a height-for-age 90% or more (p<0.05). (65)In rural Ghana, case fatality odds were 2.5 times (95% CI: 1.3-5.1) significantly higher in underweight children.(26) The authors of that study note, however, that case fatality estimates may have been overestimated as measles-related deaths were identified through informal reports.(26)

In 2008, another retrospective study also found that declining malnutrition rates among under-fives were associated with declining measles case-fatality rates. (66) Finally, a 2021 study noted that hospitalized children admitted with measles complications were 5.23 times more likely to die (p<0.0001) if they were stunted.(72)

4 community-based studies concluded that mortality from measles-associated illness was not related to undernutrition. (68, 69, 73, 74) Instead, factors such as overcrowding (i.e. clustering of cases) and the intensity of exposure to the disease were more important determinants of measles mortality. (68) (73) (74)

In 2009, a study looked at risk factors that could explain measles mortality rate variations in community-based trials. (71) The consensus regarding malnutrition was conflicting, and outcomes varied depending on how malnourished cases were defined, which lacked uniformity. (71)

In 2017, another study concluded that malnutrition, assessed in the community by comparing photos of normal and malnourished children, was not significantly associated with an increased risk of measles-related mortality.(60)

### Malnutrition and measles-related morbidity

#### Vitamin A deficiency and therapeutic supplementation

In 2017, a position paper published by the WHO indicated that vitamin A deficiency was associated with delayed recovery and a higher risk of measles-related complications.(75) Most recently, a 2022 review highlighted that vitamin A deficiency was commonly observed during measles and that this condition was associated with greater measles-related ocular morbidities and disease severity. (76) Another study in Ethiopia found that measles was one of the main, preventable underlying causes for severe visual impairment among 13.5% to 25% of blind students aged 6 years to 25 years.(77) In 2019, a review found that serum retinol levels dropped in children affected with measles, and that more important decreases were observed during severe episodes. (78)

In Zambia, acute measles patients who were vitamin A-deficient were 2.5 times (95% IC: 1.1 – 5.7) more likely to suffer from unresolved pneumonia (i.e. infected children with pneumonia at baseline which persisted after 2 weeks). (79) However, vitamin A supplementation could improve those odds. Vitamin-A deficient children who received supplementation as a single oral dose of 210 μmol retinol as retinyl esters were 80% less likely to suffer from unresolved pneumonia (OR = 0.2; 95% IC: 0.05 – 0.71). (79)

In 2016, a review of RCTs on the use of vitamin A supplementation upon hospital admission to prevent measles-related ocular morbidities concluded that evidence current to 2015 was insufficient to support the benefit of vitamin A to prevent blindness.(80) In the United States, a 2020 hospital-based study examined the median length of hospital stay among measles patients who received vitamin A and those who did not receive any doses. The study found no significant differences between the two groups, but it is notable that none of the patients had received the amount of therapeutic vitamin A currently recommended for their respective age groups.(81) In addition, laboratory assessment of vitamin A status prior to receiving supplementation was only conducted for 2 out of 142 patients (0.1%).(81) It is possible that some of the patients did not have deficient vitamin A levels, which may have reduced the impact of therapeutic vitamin A supplementation.

In 2021, another study in high-income countries found that 2 doses of oil-based vitamin A provided to children hospitalized with measles did not reduce the risk of measles-related morbidities.(82)

#### Vitamin A preventative supplementation

In India, measles complications and case-fatality rates were higher among children who did not receive routine, preventative vitamin A supplementation (p < 0.05). (83) However, the dosage supplemented was not specified.

#### Zinc therapeutic supplementation

In 2001, a study reported on an unpublished RCT which investigated zinc supplementation therapy for children hospitalized with measles and pneumonia. (54) The authors found that, after receiving 40 mg of zinc daily for 5 days, the clinical course of measles did not differ between the cases (i.e. zinc supplemented) and the control groups (i.e. placebo). (54) In 2017, a systematic Cochrane review also only returned that one randomised trial. (84) The reviewers concluded that the quality of the current evidence was very low, and that further research was necessary to determine whether zinc supplementation provided any benefits or harms for measles treatment.(84)

#### Undernutrition

In 1985, a review concluded that overcrowding was more likely to be a primary predisposing factor for severe measles than protein-energy malnutrition. (85) Crowded housing led to more secondary cases, which had more complications and a higher mortality rate than index cases. (85) The authors hypothesized that overcrowded, high-density settings led to a higher initial viral load in the infected and secondary cases then absorbed a higher amount of virus.(85) In 1986, another study found that the duration of measles symptoms (i.e. fever, rash and cough), was not significantly different across children with different levels of malnutrition. (86) In addition, no significant difference was found in the development of measles-related complications (i.e. lower respiratory infections, diarrhea) and hospitalization due to the severity of measles between well-nourished and malnourished children.(86) In Mauritania, however, factors of poor prognosis for measles patients included malnutrition (p = 0.0059). (87) Moreover, a hospital-based cohort study found that measles was a preceding precipitating factor in 30.7% of severe protein-energy malnutrition cases (i.e. kwashiorkor and marasmus). (88)

In 1979, a study found that malnourished children (i.e. those with marasmus and marasmic kwashiorkor) infected with measles were more prone to develop severe ulcerative herpes during the course of the illness (p<0.001). Mouth ulcers took on average two to three weeks to heal and, due to the pain discouraging oral intake, malnourished children lost further weight by the time this complication was resolved.(22) In 1981, another study had more conflicting findings: in non-hospitalized children, malnourished measles cases were significantly more susceptible (p<0.001) to herpes simplex virus infections than non-malnourished cases. However, this association was not observed in hospitalized, severe measles cases.(89)

Finally, in 2008, a survey found that measles-related complications were significantly higher (p < 0.001) among severely undernourished children.(27) Complications included diarrhea, pneumonia, ear infection or any other complications that led to hospitalization.(27)

#### Immunosuppression as post-measles complications

In 1971, a case-control study based on necropsies found that measles may depress cell-mediated immunity, and that a synergistic effect of measles and protein-calorie malnutrition could compound measles’ morbidity. (90) In 1973, a study concluded that immunosuppression observed during measles may be due to a direct effect of the virus rather than one mediated by malnutrition, but that severe protein-calorie malnutrition could still likely impair cell-mediated immunity through different synergistic pathways. (20) In 1980, peripheral blood mononuclear cells from malnourished and non-malnourished children were infected in vitro with measles virus. None of these children had a history of measles and were previously negative for the presence of measles-specific antibody. The study found that the subsequent viral load in the peripheral blood mononuclear cells of malnourished children was significantly greater (p<0.05).(91) In addition, the study found that cell-mediated and humoral immune response to measles infection was apparently well-preserved in a state of malnutrition: there was no significant difference between malnourished and non-malnourished children in the serum levels of antibodies produced and in the body’s ability to kill measles-infected cells.(91) Another study in 1986 found that in children infected with measles, the degree of immunosuppression was not significantly different between malnourished and well-nourished children. Cell-mediated immune response to mitogen-induced T-cell proliferation was low in all measles cases irrespective of nutritional status.(86) In 1987, a study found no significant decrease in the total white blood cell or monocyte counts between the acute, symptomatic phase and the convalescent phases of measles. (28) However, total T-cell counts were significantly lower during acute symptomatic measles (p<0.001).(28) In addition, compared to well nourished children under five years old, those who were malnourished had lower T-cell counts during acute measles (p<0.025).(28)

Another study in 2002 hypothesized that changes in lymphocyte levels during infection may contribute to measles-related immunosuppression.(92) The authors examined children hospitalized with measles and found that wasted children had lower lymphocyte counts than those who were not wasted. (92) However, lymphocyte counts did not predict disease severity. (92) In addition, stunted children were observed with higher lymphocyte levels than non-stunted ones, however the mechanisms behind these changes and their importance were unclear. (92)

In 2004, a study examined the effects of acute measles in malnourished children in Nigeria, comparing measles cases enrolled within 1 week of a generalized maculopapular rash onset and non-measles controls. (93) In cases, plasma interleukin (IL) – 12 levels were significantly lower and plasma cortisol was significantly higher. (93) IL-12 is a type of cytokine that is a key regulator of cell-mediated immune responses. (93) Cortisol, as a glucocorticoid, can suppress the synthesis and release of IL-12. (93) The authors hypothesized that impaired hepatic synthesis of a binding protein for cortisol was the cause of the marked hypercortisolemia observed in underweight, wasted and measles cases. (93)

### Findings on the potential pathways behind the association between measles and malnutrition

#### Measles predisposing to vitamin A deficiency

Vitamin A is one of the key micronutrients essential for a well-regulated immune system. In the body, its three active forms are retinol, retinal and retinoic acid. (94) Most of the mechanisms through which vitamin A modulates the immune response are driven by retinoic acid. (94) However, the form that circulates in the blood is mainly retinol. (32) Retinol is released from the liver, where most of total body vitamin A is stored, in response to tissue demand. (32) It is then converted into retinoic acid as needed. When severely depleted (<0.07 umol/L) or extremely high (> 1.05 umol/L), serum retinol is used as an indicator of hepatic vitamin A levels. (32)

None of the studies reviewed investigated how vitamin A deficiency may predispose to measles infection or to the development of measles complications. All articles examined how measles potentially instigated vitamin A deficiency.

Four studies found that hyporetinemia (i.e. serum retinol < 0.07 umol/L) occurred during acute measles episodes or was associated with measles infection. (56, 93, 95, 96)

In one review, the authors hypothesized that measles may impede the release of retinol to peripheral tissues. (56) As previously mentioned, hyporetinemia may reflect depletion of liver stores and total body vitamin A deficiency. (56). Among measles patients, however, hyporetinemia occurred even when hepatic vitamin A stores were adequate, as well as in communities where vitamin A deficiency was not prevalent. (56) In addition, following measles recovery, serum retinol rose back to normal levels without any vitamin A supplementation. (56) Another community study also noted that low serum retinol was triggered independently of the original vitamin A status. (95) Suggested causal mechanisms included an increased demand for the micronutrient while availability is decreased due to lower intake or malabsorption. (95) In Nigeria, a case-control study indicated that cytokines, released as part of the immune response, may have decreased the production of retinol-binding protein and transthyretin. (93) This would then have triggered a drop in serum retinol. (93)

### Impact of nutritional status on measles through the immune system

Two studies looked at mechanisms potentially involving the effect of nutritional status on the immune system’s response to measles.

In 1977, a hospital-based study identified malnutrition as a factor in measles persistence, defined as evidence of the virus being present (i.e. giant cells in nasal secretions and/or formed through lymphocyte response to phytohemagglutinin) six to thirteen days after onset of rash. (21) Persistent measles was found in 40% (12/30) of malnourished children post-measles and in none of the well-nourished ones (0/25). (21) In addition, compared to controls, malnourished cases had significantly blunted immune responses to tested antigens (i.e. Candida, measles). (21) It was hypothesized that malnutrition may have depressed cell-mediated immunity, prolonging the measles episode, which then further impairs the immune system and the malnourished state. (21)

In a review of clinical trials, measles led to depressed CD4 T-cells numbers. (96) CD4 T-cells can be general indicators of immune status. Measles cases who received vitamin A supplementation, however, had a significant reductions in measles morbidity and significant increases in lymphocyte levels and measles antibody concentrations. (96) Vitamin A was therefore hypothesized to improve measles prognosis by inducing improvements in CD4 T-cells numbers. (96)

### Gut barrier

In 1975, a hospital-study looked at malnourished measles cases compared to non-measles controls with moderate to severe kwashiorkor. (97) The authors found that cases patients with measles may be associated with protein-losing enteropathy. (97)

Another study also noted that gastrointestinal protein loss was significantly higher during an acute measles episode, as opposed to after recovery. (98) In that sample, all the children studied were underweight and had diarrhea. (98) In addition, a significantly higher level of gut malabsorption was observed in children with measles enteritis during an acute measles attack, indicating that acute measles may contribute to malabsorption in the proximal small intestine. (98)Faecal protein loss, in combination with malabsorption, may be a significant contributors to the development of malnutrition after measles. (98)

## Discussion

This review points to a positive correlation between vitamin A deficiency and greater incident measles case reporting, measles-related complications, and morbidity, as well as between undernutrition and incident measles reporting, as well as measles mortality. There is also evidence for a negative correlation between preventative vitamin A supplementation and measles incidence, and between therapeutic vitamin A supplementation and measles-related mortality.

### Estimating the impact of malnutrition on measles incidence

Among studies which investigated measles incidence, 31% (4/13) relied on clinical signs and symptoms, 15% (2/13) on reported cases in hospital records, and 15% (2/13) on serological results from national surveillance data to recruit measles cases. 38% (5/13) did not specify how a case was defined.

The term “incidence” throughout this manuscript is used as per the reviewed studies’ original terminology. However, while those studies aimed to evaluate the rate at which new measles cases occurred in relation to the risk factors investigated (e.g., malnutrition), there are concerns around the use of that term as incident case data may be under-reported and not accurately represent the occurrence of new measles cases over the period of time studied.

The use of clinical signs and symptoms implies that a measles case may have been more likely to be identified, especially through informal reporting, once symptoms are overt or severe (i.e., acute measles episodes). Likewise, hospital records may suggest that the disease had progressed to a point where hospitalization was required. Of the two studies which used serological confirmation, one was conducted during an outbreak. Without systematic screening, it is likely that a number of new cases may have been omitted. As such, the reported use of incidence data may not be entirely accurate and underestimated in high-risk regions. This review interprets the findings of studies on measles incidence as an impact on the likelihood of acute measles reporting rather than an impact on actual incidence.

### Vitamin A

This review shows that a well-established association exists between vitamin A deficiency and increased likelihood of incident measles case reporting, though few studies quantified the extent of that relationship. (35-37, 41-43) In addition, two meta-analyses concluded that routine vitamin A supplementation can lower reported measles incidence by 50%. (36, 37)

The evidence on the association between vitamin A deficiency, preventative vitamin A supplementation and measles mortality was more conflicting, but methodologically it is often challenging to estimate impacts on measles mortality because the identification of deaths from measles is challenging in many countries due to limitations of vital statistics, incomplete reporting and laboratory investigation of cause of death. All studies on preventative vitamin A supplementation were in community settings, where data reporting is not systematic and may be more difficult. On the other hand, high-quality studies show that therapeutic vitamin A supplementation, given in two doses of 200,000 IU, can significantly reduce measles-related mortality. In that dosage, vitamin A therapy was particularly impactful for children under 2 years old. In addition, the observed benefits of this supplementation can be greater in hospitalized children and in those who live in geographical areas with high case-fatality rates.(52)

There is also evidence of vitamin A deficiency being associated with greater measles-related complications (i.e., ocular morbidities, pneumonia), disease severity and delayed recovery.(79) Of note, however, is that those studies used serum retinol as an indicator for vitamin A deficiency. In young children, a multi-country meta-analysis (i.e., the BRINDA project) found that retinol was statistically, negatively correlated with inflammation, and that an inflammatory state could overestimate vitamin A deficiency. (99) Inflammation is triggered as part of the immune system’s reaction to harm, such as infections. When adjusted for inflammation, vitamin A deficiency prevalence dropped by − 22.1% to − 6.0%. (99) The studies reviewed did not take into account the effects of inflammation on vitamin A biomarker concentrations. If the studies reviewed had adjusted their vitamin A deficiency rates, there is a possibility that the strength of the association observed may have been weaker.

The evidence on vitamin A supplementation, both therapeutic and preventative, and measles morbidity in this review was more limited, likely due to the timeframe of the search query relating to vitamin A being restricted to 2018 to 2022. Only 4 out of the 67 studies reviewed examined therapeutic vitamin A supplementation and measles-related morbidities, and a single study reported on preventative vitamin A supplementation. Lack of routine vitamin A supplementation significantly increased measles complications, though this study did not specify whether supplementation was for previously vitaminA deficient children. (83) While three studies found that vitamin A therapy led to no significant impact on reducing measles-related morbidities, one of them highlighted that the vitamin A doses provided in their study were lower than the current recommended regimen, which may lessen the predicted effect size.

In terms of potential mechanistic pathways, historical studies seem to concur with the BRINDA project: hyporetinemia among children with measles infections may not necessarily correlate with depleted liver stores. (99) In a community trial, hyporetinemia was triggered independently of vitamin A status. (95) Another study hypothesized that measles may impede the release of vitamin A from liver stores as hyporetinemia occurred despite vitamin A stores being adequate. (56) The release of cytokines, as part of the immune inflammatory response to infections, may inhibit the production of retinol-binding protein and transthyretin. (93) Both of those transport proteins are responsible for carrying retinol from hepatic stores to target tissues, and this mechanism could explain why infections trigger hyporetinemia. Measles may increase the demand in vitamin A while body availability is decreased. (95) This would concur with previous reviews on vitamin A and infections, which found that systemic vitamin A deficiency was triggered as a result of infection-induced anorexia and malabsorption. (100)

The data on the potential pathways behind measles and vitamin A deficiency remain very limited. None of the studies reviewed looked at the potential pathways through which vitamin A supplementation could decrease measles incidence and mortality. Only one study found that vitamin A supplementation increased lymphocyte levels and measles antibody concentrations, thus reducing morbidity risk. (96)

### Undernutrition

All the studies in this review consistently reported an association between undernutrition and the increased likelihood of incident cases reporting. In all 5 studies, undernutrition significantly increased reported measles incidence by 1.7 to 24.9 times (OR: 1.7, 3.0, 3.0, 3.4, 24.9). (23, 44-46, 48) 3 studies also found that measles was an important contributing factor to malnutrition, and that it could increase the risk of malnutrition threefold. (42, 47, 48)

The evidence for an association between malnutrition and measles mortality is less consistent, though 6 out of 10 studies found a positive correlation between malnutrition and measles mortality. (62) (63) (64) (65–67) 2 of those studies found that malnutrition increased the risk of dying from measles by 1.7 and 7.6. times (62, 65) However, the estimate of 7.6 was imprecise, with a wide confidence interval from 1.3 to 44.3. (62) Reviews of community studies, on the other hand, suggested no significant association between measles mortality and malnutrition. (68, 70, 71) Wolfson et al., (2009), in particular, highlighted that those outcomes varied widely depending on the definition of malnutrition used in each study trial. Heterogeneity in malnutrition nomenclature was also noted in this review. (71)

Few studies examined the relationship between measles morbidity and malnutrition, and results among them varied. One review of community studies found that overcrowding was more likely to be a predisposing factor to severe measles than malnutrition, (85) while 2 hospital-based studies found that malnutrition and measles were precipitating factors for each other’s morbidity. (87, 88)

All studies on malnutrition and measles were observational, likely due to ethical challenges in exploring this topic with an experimental design. A randomized controlled trial cannot be conducted by purposefully exposing participants to a harmful variable such as malnutrition or measles. Observational studies, however, can be more vulnerable to biases and confounding factors, which can weaken the strength of inferences that we can draw from them. The heterogeneity of malnutrition and measles cases definitions across studies also makes estimating a combined effect size challenging and open to potential information biases (i.e. classification errors). Some of the outdated definitions, in particular, are more prone to those systematic biases and misclassification. In older studies, stunting and wasting were often classified as standalone cases. However, those two conditions share several common determinants, often leading to concurrent stunting and wasting. (101) Wasting, in particular, is associated with increased risk of stunting. (101) In an analysis of datasets from 84 countries, 37.5% of wasted cases were also concurrently stunted. (102) It is thus difficult to isolate the effects of stunting and wasting, independently, on measles. Underweight was more recently added as a composite indicator to account for cases afflicted with both stunting and wasting, however it was not in use in historical studies. Likewise, historical studies did not employ the standardized WHO classification for measles cases, and signs and symptoms used during clinical assessment were widely variable. In addition, the more recent studies do not consistently rely on the more rigorous serological testing to confirm measles cases. Almost a third of the studies reviewed identify measles cases through reported signs and symptoms, which is more prone to erroneous identification. Half of the studies did not specify how measles cases were defined.

Finally, few studies detailed the severity of malnutrition cases. Yet, the trend in which hospital studies have stronger associations than community trials may suggest that the degree of malnutrition is an underlying factor. Severely malnourished cases are more likely to be hospitalized and to be included in those studies than mild cases.

The synergy between infections and malnutrition has often been documented in the literature: chronic or recurrent infections exacerbate malnutrition, and malnutrition impairs host defense and increases susceptibility to infections (103–106) However, the causal pathways leading to that association are still inadequately understood and there is a lack of strong studies able to characterize the extent to which malnutrition contributes to infections (105). This review points to similar findings with regards to potential bidirectional relationship between measles and malnutrition. The number of studies which explored the causal mechanisms behind their association is limited (6).

### Potential pathways

Among purported mechanisms, the three main ones involve an impaired immune system, impaired gut health and gut-lung axis and/or metabolic changes.

#### Immune system and cytokines

This review suggests that malnutrition and measles may both trigger immunosuppressive pathways.

One hospital trial found that malnourished children had significantly blunted immune responses to antigens of measles and Candida yeast, a skin infection. (21) Findings from a systematic review in 2014 seem to concur, noting that undernourished children exhibit atrophy of lymphoid tissue and of the thymus gland. (107) One of the lymphatic system’s roles is to produce immune cells while the thymus develops T-cells, white blood cells responsible for recognizing antigens and fighting off infections as part of the adaptative immune response. Studies have also noted that children with kwashiorkor and marasmus produce fewer total counts of acute-phase proteins and pro-inflammatory cytokines. (103) Those signaling molecules are the body’s initial response to infection or tissue damage and are needed to activate the receptors to produce an immune reaction. (103) In addition, undernutrition can shorten the life span of monocytes, white blood cells involved in the recognition and destruction of harmful organisms. (103) Regarding measles and cell-mediated immunity, one study identified that measles decreased the levels of circulating IL-12 cytokines in malnourished children. (93) As IL-12 is produced by cells such as monocytes, this could potentially be one of the pathways that mutually reinforces the synergy between malnutrition and measles. (93)

Previous studies have also found that measles infection had an impact on the lymphatic system, and that it was associated with lower levels of circulating lymphocytes and with their depletion in lymphoid tissues. (108) However, while lymphocytes levels normalized after the resolution of measles symptoms, immune suppression persisted for several weeks to years after the initial infection. (108, 109) Those findings suggest that measles may be triggering disease-specific immune mechanisms. (108) A 2019 study highlighted that the measles virus infects key immune cells, including memory cells. (109) Following infection, immunological memory seems to be restructured, leading to a loss of memory cells from previous infections and to an impaired bone marrow. (109) The bone marrow normally produces naïve B-cells, which are meant to become memory cells that secrete antibodies after being exposed to an antigen. (109) Under normal physiological conditions, constant repopulation of naïve B-cells ensures that the body can maintain a diverse antibody repertoire. (109) Immune diversity, however, was severely diminished in 10% of measles cases. (109) In another study, measles pruned back 11% to 73% of pre-existing antibody repertoires. (110) Through this mechanism, measles likely causes a reduced recognition of infectious pathogens and an increased susceptibility to infections, including diseases against which a child was previously vaccinated.

Current understandings indicate that both measles and malnutrition suppress the immune system. Malnutrition can be a factor in measles persistence, akin to the effect of measles on the lymphatic system. (21) It is possible that co-occurring conditions in an individual might magnify the effects observed through those immune pathways and exacerbate malnutrition and/or measles outcomes. Further studies to confirm this hypothesis have to be conducted.

#### Gut health and gut-lung axis

Studies on gut health demonstrate that the gut interacts closely with the immune system. The surface of the gut is an immune sensor that detects pathogens, and the intestinal wall regulates the passage of viable bacteria from the gastrointestinal tract to other sites. (104) In addition, recent understandings on the gut’s microbiome suggest that it is highly involved with immune homeostasis. (104) In a 2018 review, dysbiosis (i.e. imbalance in the gut’s microbiome) was found to be a precursor to infection. (104)

Two historical studies also found that malnourished measles cases, as opposed to non-measles malnourished controls, were associated with protein-losing enteropathy and malabsorption. Both outcomes can precipitate an already nutritionally deficient state, in addition to worsening an already vulnerable immune status. (97, 98)

In addition, malnutrition was found to be associated with disturbances to the integrity of the intestinal mucosa, gut-barrier functions and gut microbial communities. (103, 104) A systematic review found that malnourished children had atrophied intestinal walls, which would likely have increased intestinal permeability and predisposed those children to bacterial translocation from the gut into the bloodstream. (107) Atrophied intestinal mucosa would also have a reduced surface area for absorption. Children with an impaired gut likely do not absorb the quantity of nutrients required to sustain an optimal nutritional status. (104) These findings may concur with the ones from historical studies, though they may also point to a role for the degree of malnutrition.

None of the studies reviewed explored how measles might lead to those consequences. However, growing evidence reinforces the concept of a gut-lung axis, in which respiratory infections may be able to impact gut health. (111) In response to infections, recent research suggests that immune cells and metabolites use the mesenteric lymphatic system to migrate from the gut to the lungs and modulate lung immune response. (111, 112) The presence of certain types of gut bacteria is essential to produce those cells at the gut site, and dysbiosis might displace them. (112) In addition, studies have documented events of lung diseases triggering increases in the gut’s bacterial load, changing its composition. (112) The gut-lung axis could potentially be an alternative pathway through which measles and malnutrition reinforce their synergistic relationship, though further evidence is required to refine this understanding.

### Metabolic factors

While this review did not identify any potential metabolic factors in the relationship between measles and malnutrition, infections are known to impair child growth via several pathways: increased leptin secretion, increased energy expenditure, limited nutrient availability and uptake, and disruption in the digestive systems’ regulation. (113)

For instance, sustained release of cytokines in response to chronic infections causes an increase in leptin secretion, a hormone which suppresses the appetite. (106, 113) Persistent measles identified in malnourished children could therefore compound undernutrition in those children. This acute-phase response to infection also tends to lead to a negative energy balance. (103) As part of their role as signaling molecules, cytokines can mediate the induction of fever, and energy expenditure is increased by 7 to 11% for each unit (degrees Celsius) increase in fever. (103) In people infected with measles, fever usually ranges from 39°C to 40.5°C and lasts 4 to 7 days. (114) If there are complications, fever may persist ever longer. (114) In addition, pro-inflammatory cytokines can inhibit the process by which bone tissue and the growing skeleton are formed. (106)

Activating the immune system may also limit the availability of certain key nutrients (e.g. iron, vitamin A, zinc), diverting them away from responding to nutritional needs. (113)Finally, older studies of severely malnourished children have also found that their gastric acid production was greatly reduced, increasing their susceptibility to infection. (103)

### Limitations

A scoping review was conducted as this paper aims to investigate the current state of knowledge on measles and malnutrition, and to map the evidence gaps. As such, this manuscript is not a systematic review, however it represents an overall assessment of the current body of evidence. Nine studies were also excluded despite reporting a measure of association as the effect of measles on malnutrition, or the effect of malnutrition on measles, could not be isolated from other variables (i.e. exclusion criteria). Search queries including terms related to “vitamin A”, “serum retinol” and “hyporetinemia” were limited to studies published 2018 thereafter. Moreover, the pool of studies that examined direct mechanistic pathways between malnutrition and measles was very small. There were also no consistent definitions for malnutrition and measles cases, which led to difficulties in estimating a cumulative effect size for associations between the nutritional status and measles.

All study designs on undernutrition were descriptive or observational, which reduced their internal validity. Moreover, few studies examining the impact of measles on undernutrition controlled for potential confounding factors such as maternal nutrition, postnatal diet, breastfeeding practices, maternal height, parental education, household assets and micronutrient deficiencies such as zinc and iron. Those factors are determinants of childhood malnutrition. (101) Some studies investigating the effect of malnutrition on measles did not control for vaccination status and many did not specify the epidemiological setting (e.g., endemic, outbreak). Both variables could modify the observed rates of measles incidence, severity and mortality. In addition, out of the studies that examined the impact of vitamin A supplementation on measles, few specified whether prior vitamin A status of the children receiving supplementation was investigated. Pre-existing vitamin A deficiency could have been a modifying factor in the results observed. The supplementation regimens varied widely, and some had unspecified dosage. Moreover, in studies that assessed vitamin A status, inflammation was seldom taken into account, despite it being able to cause overestimations in vitamin A deficiency. Due to scope and time restrictions, we did not include studies whose main topic was on noma, a disease that has often been noted in measles and malnutrition settings. While some studies accounted for measles-related gastrointestinal complications, none of them examined noma as a secondary variable in the measles-malnutrition relationship either.

Finally, this review was limited to studies published in English and in French. S6 Table summarizes the methodological limitations of this review.

### Evidence Gaps

The current state of knowledge on preventative vitamin A is strongest in relation to reported incident measles cases. Statistical evidence on the role of preventative vitamin A supplementation on measles-related deaths, however, could be improved. In terms of vitamin A therapy, high doses, provided for hospitalized, young children, and in high case-fatality rates settings, seems more likely to derive benefits from the supplementation. However, the extent to which each of those factors individually contribute to the effect size has not been investigated. Regarding vitamin A deficiency, emerging evidence throws into question the use of serum retinol as a reliable indicator of vitamin A status in a context of inflammation.

Evidence gaps remains in assessing the potential impacts of these findings on previous conclusions on vitamin A deficiency and measles.

Current evidence on measles and malnutrition relies on narrative reviews and original research reports of descriptive and observational studies, likely due to ethical challenges in designing randomized controlled trials for this topic. Across all studies reviewed, there was also high variability in the effects or outcomes being evaluated due to the use of different undernutrition definitions. Variability in underlying populations was also present, as studies encompassed settings in hospitals, communities and acute crises (i.e., conflict, famine). This lack of homogeneity may be a challenge to conduct a meta-analysis with a stronger, cumulative effect size.

Little data was available to shed light on the potential pathways behind malnutrition and measles. On the other hand, many studies explore the pathophysiology of measles and malnutrition separately. Further research is needed to fully explain how measles impacts the nutritional state, and vice versa.

Finally, while certain key nutrients are known to have an active role in the immune system (e.g., iron, zinc), this review identified few articles which isolated the impact of micronutrients other than vitamin A. The only studies on the effect of therapeutic zinc supplementation on measles-related morbidities concluded that the current evidence was of very low quality and that further research was necessary.

## Conclusion

The findings from this review reinforces the hypothesis of a synergistic relationship between measles and malnutrition. Measles infection is an important contributing factor to malnutrition, potentially increasing the risk of being malnourished threefold. Observational data has also consistently underlined a positive correlation between undernutrition and reported incident measles case data. Evidence on the potential impacts of malnutrition on measles mortality is weaker, however a positive correlation has been documented between both conditions. The extent to which malnutrition affects measles mortality, however, remains imprecise. Few studies examined the relationship between measles morbidity and malnutrition, and results among them were conflicting. Likewise, few studies reported on vitamin A supplementation, both therapeutic and preventative, and measles-related morbidities. It is therefore difficult to conclude on the association between these variables. However, the most recent Cochrane meta-analysis concluded that vitamin A supplementation was associated with a clinically meaningful reduction in childhood morbidity, highlighting its benefits for non-specific morbidities.(38)

All studies reviewed found an association between vitamin A deficiency and the increased likelihood of incident measles reporting, measles-related complications and/or death. Two doses of 200,000 IU therapeutic vitamin A could significantly reduce the odds of measles-related mortality. There is also robust evidence that supports the use of preventative vitamin A campaigns to reduce measles incidence. Moreover, though the two most recent reviews of randomized trials found that preventative vitamin A had non-significant effects on mortality specifically due to measles, there is high-quality evidence for vitamin A supplementation reducing the risk of all-cause infantile mortality by 12% to 24%.(37, 115) Routine provision of vitamin A supplements therefore remains highly beneficial, especially in areas where vitamin A deficiency is a public health concern, and where measles is endemic.

Incorporating a nutrition strategy into public health frameworks aiming to prevent and combat measles outbreaks may therefore be a necessity. The available evidence for a bidirectional relationship between malnutrition and measles disease supports the use of integrated platforms to simultaneously provide both vaccination and nutritional services, particularly in globally underserved communities where multi-dimensional depravations across health and nutrition sectors are likely to occur. Current WHO guidelines recommend bi-annual delivery of preventative vitamin A supplements to children 6 to 59 months old, integrated into public health initiatives such as national measles vaccination campaigns. (115)

The potential pathways between malnutrition and measles remain poorly defined, and the number of well-conducted studies on undernutrition and measles is limited. Much of the research is dated; the field of nutritional research has made many advances in the interim. Further research on the specific causal mechanisms underlying measles and malnutrition is needed. Insight on this evidence gap may allow us to refine better pathophysiological targets for both therapeutic and preventive interventions. Additional studies with the use of a standardized, consistent approach to defining malnutrition would also be important to ensure the comparability of outcomes between different studies and trials. This would facilitate the determination of a cumulative effect size and allow for stronger inferences of findings to policy making.

## Supporting information

S1 File. Sample search strategy in Pubmed

S1 Table. Characteristics of included studies on malnutrition and measles

Preferred Reporting Items for Systematic reviews and Meta-Analyses extension for Scoping Reviews (PRISMA-ScR) Checklist

S2 Table. Distribution of different malnutrition nomenclature across studies

S3 Table. Definitions of variables

S4 Table. Characteristics of vitamin A deficiency studies

S5 Table. Characteristics of vitamin A supplementation studies

S6 Table. Summary of methodological limitations in the existing evidence base

## Data Availability

All data produced in the present work are contained in the manuscript.

## Acknowledgements

This scoping review was initiated as part of a remunerated internship with the Early Childhood Nutrition Unit of the United Nations Children’s Fund (UNICEF) and was completed within a research contract funded by the World Health Organization (WHO).

The final research paper is the result of the generous support and sharing of insight from many individuals who provided their time and expertise throughout its development, in particular: Natasha S Crowcroft (Department of Immunisation, Vaccines, and Biologicals, World Health Organization, Geneva, Switzerland) and Andreas Hasman (UNICEF Programme Division, New York, New York, USA), for their mentorship, feedback on draft versions of the manuscript, and guidance in structuring this review and formulating its overarching goals, refining the search methodology and coordinating this project’s execution; Tracey S Goodman (Department of Immunization, Vaccines and Biologicals, World Health Organization, Geneva, Switzerland), for her guidance, incredible patience in responding to questions and inquiries, and feedback on the manuscript; and Kim Mulholland (Infection and Immunity Theme, New Vaccines group, Murdoch Children’s Research Institute, Melbourne, Australia; Department of Paediatrics, University of Melbourne, Melbourne, Australia; Department of Infectious Disease Epidemiology, London School of Hygiene and Tropical Medicine, London, United Kingdom) and Robert Bandsma (Centre for Global Child Health, The Hospital for Sick Children, Toronto, Ontario, Canada; Department of Nutritional Sciences, Faculty of Medicine, University of Toronto, Toronto, Canada), for their critical input on the key pieces of scientific literature and important areas of investigation to focus on.

## Supporting information

**S1 Table.** Characteristics of included studies on malnutrition and measles.

**S2 Table.** Distribution of different malnutrition nomenclature across studies.

**S3 Table.** Definitions of variables.

**S4 Table.** Characteristics of vitamin A deficiency studies.

**S5 Table.** Characteristics of vitamin A supplementation studies.

**S6 Table.** Summary of methodological limitations in the existing evidence base.

## Notes

### Competing Interest Statement

ICTT had a remunerated internship with the Early Childhood Nutrition Unit of the
United Nations Children's Fund (UNICEF) to initiate this study, and a research contract
with the World Health Organization (WHO) to complete and publish it.
CG and AI contributed to the design of the study and its overarching goals, and to the
research and analysis methodology.
CG, POC and AI contributed to the review of the manuscript.

