## Supplementary material for "A scoping review on the associations and potential pathways between malnutrition and measles": S1 File. Sample search strategy in Pubmed

| **Database:** Pubmed | | | |
| --- | --- | --- | --- |
| **Filters:** English and French; full text available | | | |
| **#** | **Query** | **Results without filters** | **Results with filters** |
| S1 | “measles” [All fields] | 30 990 | 9 322 |
| S2 | “malnutrition” [All fields] | 160 481 | 37 471 |
| S3 | "malnutrition " OR "malnourished" OR "maln*” | 63 320 | 18 898 |
| S4 | "malnutrition " OR "malnourished" OR "maln *" OR "undernutrition" OR "undernourish*" | 68 866 | 21 116 |
| S5 | S1 AND S2 | 708 | 400 |
| S6 | S1 AND S3 | 578 | 316 |
| S3 | S1 AND S4 | 613 | 335 |
| Example | All fields = "measles" AND ("malnutrition " OR "malnourished" OR "maln *" OR "undernutrition" OR "undernourish*") | | |

**S1 File.** Sample search strategy in Pubmed (as of Feb 1^st^ 2022)
