## Supplementary material for "A scoping review on the associations and potential pathways between malnutrition and measles": S1 Table. Characteristics of included studies on malnutrition and measles

| **S1 Table: Characteristics of included studies on malnutrition and measles** | | | | | | | |
| --- | --- | --- | --- | --- | --- | --- | --- |
| **Authors** | **Date** | **Type of study** | **Measles aspect** | **Setting** | **Variable of interest** | **Outcome** | **Measure of association** |
| Hübschen et al. | 2022 | Review | Morbidity | Not specified | Therapeutic vitamin A supplementation and vitamin A deficiency | Measles morbidity | Not available |
| Wang et al. | 2022 | Review | Mortality | Not specified | Wasting, underweight, stunting, vitamin A deficiency | Measles mortality | Not available |
| Nassar et al. | 2021 | Analytical study: observational | Incidence | Community | Acute malnutrition / Wasting | Measles incidence | OR: 24.9  (95% CI: 1.9-329.6) |
| Donadel et al. | 2021 | Analytical study: observational | Incidence | Hospital | Malnutrition | Measles incidence | OR: 3.4  (95% CI: 1.1, 9.9) |
| Aurangzeb et al. | 2021 | Analytical study: observational | Mortality | Hospital | Malnutrition | Measles mortality | 1. Underweight OR: 2.93 (CI: 1.44-5.93, p = 0.003)  2. Stunting OR: 6.8 (CI: 3.24-14.26, p<0.0001) |
| Lo Vecchio et al. | 2021 | Analytical study: observational | Morbidity | Hospital | Therapeutic vitamin A supplementation | Measles severity / morbidity | RR: 1.33 (95% CI: 0.59-2.96) |
| Lowang et al. | 2021 | Analytical study: observational | Incidence | Community | Preventative vitamin A supplementation | Measles incidence | RR: 0.23 (95% CI: 0.08-0.66) |
| Domai et al. | 2021 | Analytical study: observational | Mortality | Hospital | Therapeutic vitamin A supplementation | Mortality | OR: 1.93 (95% CI: 1.31-2.85) |
| Hester GZ, Nickel AJ, Stinchfield PA, Spaulding AB. | 2020 | Analytical study: observational | Morbidity | Hospital | Vitamin A supplementation | Measles severity / morbidity | RD: no significant differences |
| Dureab et al. | 2019 | Analytical study: observational | Incidence | Community | Acute malnutrition / Wasting | Measles incidence | OR: 3.2 (p<0.000) |
| Strebel PM, Orenstein WA. | 2019 | Review | Mortality, morbidity | Not specified | Therapeutic vitamin A supplementation and vitamin A deficiency | Measles mortality and morbidity | Not available |
| Ntirampeba et al. | 2018 | Analytical study: observational | Incidence | Community | Malnutrition | Measles incidence | Not available |
| Ajibola et al. | 2017 | Review | Morbidity | Hospital | Zinc supplementation | Measles morbidity | Not available |
| Mahmud et al. | 2017 | Descriptive study | Mortality | Community | Chronic malnutrition / Stunting | Measles mortality | Not available |
| Imdad et al. | 2017 | Meta-Analysis | Incidence | Community | Vitamin A supplementation | Measles incidence | RR: 0.50  (95% CI: 0.37 to 0.67) |
| Mafigiri et al. | 2017 | Analytical study: observational | Mortality | Community | 1. Lack of vitamin A supplementation; 2. Malnutrition | Measles mortality | 1. OR: 2.3 (95% CI: 0.74-7.4)  2. OR: 2.5 (95% CI: 0.65-9.1) |
| Asferaw M, Woodruff G, Gilbert C. | 2017 | Descriptive study | Morbidity | Community | Measles, vitamin A deficiency | Measles morbidity (corneal blindness) | Not available |
| World Health Organization | 2017 | Review | Morbidity | Not specified | Vitamin A deficiency and supplementation | Measles morbidity / severity | Not available |
| Bello et al. | 2016 | Systematic review | Morbidity | Hospital, Community | Vitamin A supplementation | Measles-related ocular morbidities | Not available |
| Stevens et al. | 2015 | Analytical study: observational | Mortality | Not specified | Vitamin A deficiency, vitamin A supplementation | Measles mortality | RR: 1.26  (95% CI: 0.84 - 1.87) |
| Boushab et al. | 2015 | Descriptive study | Morbidity | Hospital | Malnutrition | Measles morbidity | Not available |
| Mahamud et al. | 2013 | Analytical study: observational | Mortality | Hospital | Acute malnutrition / Wasting | Measles mortality | OR: 7.6  (95% CI: 1.3-44.3) |
| Sachdeva et al. | 2011 | Analytical study: observational | Incidence | Community | Measles | Vitamin A deficiency | OR: 8.9 (95% CI: 6.4-12.5) |
| Mayo-Wilson et al. | 2011 | Meta-Analysis | Incidence | Not specified | Vitamin A supplementation | Measles incidence | RR: 0.50  (95% CI: 0.37 to 0.67) |
| Wolfson et al. | 2009 | Review | Potential causal pathways, mortality | Community, hospital | Malnutrition | Measles' case fatality rates | Not available |
| Saleem et al. | 2009 | Analytical study: observational | Incidence | Hospital | Measles | Malnutrition rates | OR: 3.02 (p=0.039) |
| Mishra et al. | 2008 | Analytical study: observational | Morbidity | Community | Vitamin A supplementation | Measles' complications rates | Chi-square test: 44.55, df=1, p<0.001 |
| Marufu et al. | 2008 | Analytical study: observational | Mortality | Hospital, Community | Malnutrition | Measles' case fatality rates | Slope: 0.459 (95% CI: 0.031-0.099;  R2: 0.867) |
| Mishra et al. | 2008 | Analytical study: observational | Morbidity | Community | Vitamin A supplementation | Measles' complications rates | Chi-square test: 44.55, df=1, p<0.001 |
| Schaible & Kaufmann | 2007 | Review | Potential causal pathways | Not specified | Not applicable | Not applicable | Not available |
| Chisti et al. | 2007 | Analytical study: observational | Incidence | Hospital | Measles | Rates of:  Underweight; Acute malnutrition / Wasting; Chronic malnutrition / Stunting | OR: 1.7  (95% CI: 1.3-2.4) |
| Huiming et al. | 2005 | Meta-Analysis | Mortality | Hospital, Community | Vitamin A supplementation | Measles mortality | All studies:  RR: 0.70 (95% CI: 0.42 - 1.15) Hospital-specific:  RR: 0.18 (95% CI: 0.03 to 0.61) |
| Perry & Halsey | 2004 | Review | Potential causal pathways, mortality | Not specified | Not applicable | Not applicable | Not available |
| Phillips et al. | 2004 | Analytical study: observational | Potential causal pathways | Community | Measles | Metabolic effects (e.g. hyporetinemia) | Not available |
| Caulfield et al. | 2004 | Review | Mortality | Hospital, Community | Underweight | Measles mortality | Not available |
| D'Souza RM & R D'Souza | 2002 | Meta-Analysis | Mortality | Hospital, Community | Vitamin A supplementation | Measles mortality and pneumonia-related mortality | Summary RR for 1 and 2 doses on overall mortality: 0.61 (95% CI: 0.32-1.12)  Summary RR for 2 doses on overall mortality: 0.36 (95% CI: 0.14-0.82)  Summary RR for 2 doses on pneumonia-related mortality: 0.33 (95% CI: 0.08-0.92 |
| Rosales | 2002 | Analytical study: experimental (RCT) | Morbidity, Potential causal pathways | Hospital | Vitamin A supplementation | Measles-related pneumonia | OR: 0.20  (95% CI: 0.05-0.71) |
| Ryon et al. | 2002 | Analytical study: observational | Potential causal pathways | Hospital | Malnutrition | Lymphocyte changes | Not available |
| Mahalanabis & Bhan | 2001 | Review | Mortality | Not specified | Vitamin A supplementation | Measles mortality | Not available |
| Khandait et al. | 2000 | Analytical study: observational | Incidence | Community | Measles | Vitamin A deficiency | OR: 9.26 (95% CI: 2.75-31.19) |
| Sommer | 1997 | Meta-Analysis | Mortality | Hospital, Community | Vitamin A supplementation | Measles mortality | Community:  OR: 0.5 |
| Dollimore et al. | 1997 | Analytical study: experimental (RCT) | Incidence, mortality | Community | Vitamin A supplementation | Measles incidence, measles mortality | RD: 5.3 fewer measles cases per 1,000 child-years |
| Semba | 1994 | Review | Potential causal pathways | Not specified | Vitamin A supplementation, vitamin A deficiency | Measles morbidity | Not available |
| Madhulika et al. | 1994 | Analytical study: observational | Mortality | Hospital | Vitamin A supplementation | Measles' case fatality rates | Not available |
| Fawzi et al. | 1993 | Meta-Analysis | Mortality | Hospital | Vitamin A supplementation | Measles mortality | OR: 0.39 (95% CI: 0.22 to 0.66) |
| Hussey & Klein | 1992 | Review | Potential causal pathways | Hospital | Measles | Vitamin A deficiency | Not available |
| Caballero & Rice | 1992 | Review | Potential causal pathways | Community | Measles | Vitamin A deficiency | Not available |
| Aaby | 1988 | Review | Mortality | Hospital, Community | Malnutrition | Measles mortality | Not available |
| Aaby et al. | 1988 | Analytical study: observational | Mortality | Community | Malnutrition | Measles morbidity | OR: 0.7 (95% CI: 0.3-1.6) |
| De Sole et al. | 1987 | Analytical study: observational | Incidence | Community | Vitamin A deficiency | Measles incidence | RR: 4.7 (p = 0.01) |
| Barclay et al. | 1987 | Analytical study: experimental (RCT) | Mortality | Hospital | Vitamin A supplementation | Measles mortality | Not available |
| Dagan et al. | 1987 | Descriptive study | Potential causal pathways | Hospital | Malnutrition | T lymphocyte levels | Not available |
| Bhaskaram et al. | 1986 | Analytical study: observational | Potential causal pathways | Hospital | Malnutrition | Measles morbidity | Not available |
| Aaby & Coovadia | 1985 | Review | Morbidity | Hospital | Protein-energy malnutrition (PEM) | Measles morbidity | Not available |
| Oyedeji | 1984 | Descriptive study | Morbidity | Hospital | Measles incidence and complications | Marasmus, Kwashoirkor | Not available |
| Aaby et al. | 1984 | Analytical study: observational | Mortality | Community | Malnutrition | Measles morbidity | OR: 0.7 (95% CI: 0.3-1.6) |
| Lepage | 1983 | Descriptive study | Mortality | Hospital | Malnutrition | Measles mortality | Not available |
| Koster et al. | 1981 | Descriptive study | Mortality | Community | Measles | Malnutrition | Not available |
| Orren et al. | 1981 | Analytical study: observational | Morbidity | Hospital | Malnutrition | Measles-related susceptibility to Herpes Simplex Virus Infections | Not available |
| Whittle et al. | 1980 | Analytical study: observational | Potential causal pathways | Not specified | Malnutrition | Measles viral load | Not available |
| Whittle et al. | 1979 | Analytical study: observational | Potential causal pathways | Not specified | Malnutrition | Measles viral load | Not available |
| Dossetor et al. | 1977 | Analytical study: observational | Potential causal pathways | Hospital | Kwashiorkor, Marasmus, Underweight, Marasmic kwashiorkor | Not applicable | Chi-square test: 7.1, p<0.01 |
| Dossetor & Whittle | 1975 | Descriptive study | Potential causal pathways | Hospital | Underweight, Measles-related enteritis | Not applicable | Not available |
| Axton | 1975 | Descriptive study | Potential causal pathways | Hospital | Measles, kwashiorkor | Not applicable | Not available |
| Whittle et al. | 1973 | Analytical study: observational | Potential causal pathways | Not specified | Malnutrition | Measles viral load | Not available |
| Smythe et al. | 1971 | Analytical study: observational | Potential causal pathways | Hospital | Measles, protein-calorie malnutrition | Not applicable | Not available |
| Morley | 1969 | Descriptive study | Incidence | Hospital | Measles | Acute malnutrition / Wasting | Not available |
