## Supplementary material for "A scoping review on the associations and potential pathways between malnutrition and measles": S2 Table. Distribution of different malnutrition nomenclature across studies

| **S2 Table: Distribution of different malnutrition nomenclature across studies*** | | | |
| --- | --- | --- | --- |
| **Malnutrition nomenclature** | **Number of studies** | **Timeframe in which this term was used** | **Studies** |
| Acute malnutrition | 4 | 1983-2019 | Dureab et al. (2019); Mahamud et al (2013); Saleem et al. (2009); Lepage et al (1983) |
| Protein-calorie malnutrition (PCM) | 2 | 1971-1973 | Whittle et al. (1973); Smythe et al. (1971) |
| Protein-energy malnutrition (PEM) | 4 | 1981-2007 | Schaible & Kaufmann (2007); Aaby & Coovadia (1985); Oyedeji (1984); Koster et al. (1981) |
| Kwashiorkor | 6 | 1975-1986 | Bhaskaram et al. (1986); Oyedeji (1984); Whittle et al. (1980) Dossetor (1977) Axton (1975) |
| Marasmus | 4 | 1977-1984 | Oyedeji (1984); Orren et al. (1981); Whittle et al. (1979) Dossetor (1977) |
| Marasmic kwashiorkor | 5 | 1977-1986 | Bhaskaram et al. (1986); Oyedeji (1984); Whittle et al. (1980) Whittle et al. (1979); Dossetor (1977) |
| Wasting | 7 | 1988-2022 | Wang et al. (2022); Sachdeva et al. (2011); Wolfson et al. (2009); Chisti et al. (2007); Phillips et al. (2004); Ryon et al. (2002); Aaby (1988) |
| Stunting | 7 | 1988-2022 | Wang et al. (2022); Aurangzeb et al. (2021) Sachdeva et al. (2011); Wolfson et al. (2009); Chisti et al. (2007); Ryon et al. (2002); Aaby (1988) |
| Underweight | 12 | 1975-2022 | Wang et al. (2022); Aurangzeb et al. (2021); Ntirampeba et al. (2018); Wolfson et al. (2009); Chisti et al. (2007); Phillips et al. (2004); Caulfield et al. (2004); Aaby (1988); Aaby et al. (1988); Aaby et al. (1984); Dossetor (1977); Dossetor & Whittle (1975) |
| Not specified | 15 | N/A | Wang et al. (2022); Nassar et al. (2021); Donadel et al. (2021); Mahmud et al. (2017); Mafigiri et al. (2017); Boushab et al. (2015); Wolfson et al. (2009); Marufu et al. (2008); Perry & Halsey (2004); Dollimore et al. (1997); Aaby (1988); Dagan et al. (1987); Orren et al. (1981); Morley (1969) |

*This table only presents the distribution of definitions across studies that covered undernutrition. More than one nomenclature may have been used in a given study.
