## Supplementary material for "A scoping review on the associations and potential pathways between malnutrition and measles": S3 Table. Definitions of variables

| **S3 Table: Definitions of variables** | | | | |
| --- | --- | --- | --- | --- |
| **Authors** | **Date** | **Determination of a measles case** | **Malnutrition nomenclature** | **Definition of a malnourished case** |
| Hübschen JM et al. | 2022 | Not specified | Not applicable | Not applicable |
| Wang et al. | 2022 | Not specified | Not specified | Not specified |
| Nassar et al. | 2021 | Suspected cas as per WHO | Not specified | Mid-upper arm circumference (MUAC) <115 mm |
| Donadel et al. | 2021 | Laboratory or epidemiological linkage | Not specified | Indicators not specified (clinical evaluation by a health professional) |
| Aurangzeb et al. | 2021 | Probable case as per WHO | Stunting, underweight | Weight-for-age (WAZ), Height-for-age (HAZ) |
| Lo Vecchio et al. | 2021 | Clinicals signs and symptoms & serologically confirmed | Not applicable | Not applicable |
| Lowang et al. | 2021 | Clinical signs and symptoms & serologically confirmed | Not applicable | Not applicable |
| Domai et al. | 2021 | Suspected measles as per WHO | Not applicable | Not applicable |
| Hester et al. | 2020 | Not specified | Not applicable | Not applicable |
| Dureab et al. | 2019 | Probable case (fever, rash, cough and/or conjunctivis + dx as suspected measles by healthcare) | Acute malnutrition (SAM, MAM) | Weight-for-height (WHZ), Mid-upper arm circumference (MUAC) |
| Strebel PM & WA Orenstein | 2019 | Not specified | Not applicable | Not applicable |
| Ntirampeba et al. | 2018 | Clinically-confirmed case or an epidemiological linked case as per WHO or serologically confirmed | Underweight | Weight-for-age (WAZ) z-score <-3 |
| Ajibola et al. | 2017 | Not specified | Not applicable | Not applicable |
| Mahmud et al. | 2017 | Mortality records (signs and symptoms) | Not specified | Not specified |
| Imdad et al. | 2017 | Not specified | Not applicable - Vitamin A | Not applicable - Vitamin A |
| Mafigiri et al. | 2017 | Probable and confirmed cases (IgM) as per WHO | Not specified | Weight, mid-upper arm circumference (MUAC), clinical signs and symptoms (edema), photos as proxy of normal and malnourished children |
| Asferaw et al. | 2017 | Not specified | Not applicable | Not applicable |
| World Health Organization | 2017 | As per WHO | Not specified | Not specified |
| Bello et al. | 2016 | Not specified | Not applicable - Vitamin A | Not applicable - Vitamin A |
| Stevens et al. | 2015 | Not specified | Not applicable - Vitamin A | Not applicable - Vitamin A |
| Boushab et al. | 2015 | Probable case as per WHO | Not specified | Not specified |
| Mahamud et al. | 2013 | Probable case as per WHO | Acute malnutrition | WHO 2006 standards:  Weight-for-height (WHZ) for <5 years old; edema or extreme thinness for ≥5 years old |
| Sachdeva et al. | 2011 | Not specified | Wasting, stunting | CDC 2000 standards: Weight-for-height (WHZ), Height-for-age (HAZ) z-score <-2 |
| Mayo-Wilson et al. | 2011 | Not specified | Not applicable - Vitamin A | Not applicable - Vitamin A |
| Wolfson et al. | 2009 | Varying definitions per study included in the review | Variable (not specified, underweight, wasting, stunting) | Variable (not specified, weight-for-height (WHZ), height-for-age (HAZ)) |
| Saleem et al. | 2009 | Hospital records | Severe acute malnutrition | Not specified |
| Mishra et al. | 2008 | Probable case as per WHO | Not applicable - Vitamin A | Not applicable - Vitamin A |
| Marufu et al. | 2008 | Not specified | Not specified | Indicators not specified (nutrition mortality data) |
| Mishra et al. | 2008 | Probable case as per WHO | Not applicable - Vitamin A | Not applicable - Vitamin A |
| Schaible & Kaufmann | 2007 | Not applicable | Protein-energy malnutrition (PEM) | Not specified |
| Chisti et al. | 2007 | Not specified | Underweight, wasting, stunting | Weight-for-height (WHZ), Height-for-age (HAZ), Weight-for-age (WAZ) z-score <-3 |
| Huiming et al. | 2005 | Not specified | Not applicable - Vitamin A | Not applicable - Vitamin A |
| Perry & Halsey | 2004 | Not specified | Not specified | Not specified |
| Phillips et al. | 2004 | Suspected cas as per WHO | Underweight, wasting | Weight-for-age (WAZ), Weight-for-height (WHZ) z-score <-2 |
| Caulfield et al. | 2004 | Not specified | Underweight | Weight-for-age (WAZ) |
| D'Souza RM & R D'Souza | 2002 | Not specified | Not applicable - Vitamin A | Not applicable - Vitamin A |
| Rosales | 2002 | Serologically confirmed | Not applicable - Vitamin A | Not applicable - Vitamin A |
| Ryon et al. | 2002 | Serologically confirmed | Wasting, stunting | Weight-for-height (WHZ), Height-for-age (HAZ) |
| Mahalanabis & Bhan | 2001 | Not specified | Not applicable - Vitamin A | Not applicable - Vitamin A |
| Khandait et al. | 2000 | Not specified | Not applicable - Vitamin A | Not applicable - Vitamin A |
| Sommer | 1997 | Not specified | Not applicable - Vitamin A | Not applicable - Vitamin A |
| Dollimore et al. | 1997 | Reported to have had measles in the previous 4 months | Not specified | Weight-for-age (WAZ), mid-upper arm circumference (MUAC) |
| Semba | 1994 | Not specified | Not applicable - Vitamin A | Not applicable - Vitamin A |
| Madhulika et al. | 1994 | Medical history and physical examination | Not applicable - Vitamin A | Not applicable - Vitamin A |
| Fawzi et al. | 1993 | Hospital records (hospital studies) and unspecified (community studies) | Not applicable - Vitamin A | Not applicable - Vitamin A |
| Hussey & Klein | 1992 | Not specified | Not applicable - Vitamin A | Not applicable - Vitamin A |
| Caballero & Rice | 1992 | Serologically confirmed | Not applicable - Vitamin A | Not applicable - Vitamin A |
| Aaby | 1988 | Not specified | Variable (not specified, underweight, wasting, stunting) | Weight-for-height (WHZ), Height-for-age (HAZ), Weight-for-age (WAZ), Mid-upper arm circumference (MUAC) |
| Aaby et al. | 1988 | Not specified | Underweight | Weight-for-age (WAZ) |
| De Sole et al. | 1987 | Local name and presence of other cases in the community in the same period of time | Not applicable - Vitamin A | Not applicable - Vitamin A |
| Barclay et al. | 1987 | Clinical signs and symptoms (rash) and history of prodromal disease | Not applicable - Vitamin A | Not applicable - Vitamin A |
| Dagan et al. | 1987 | Serologically confirmed | Not specified | Weight-for-age (WAZ), mid-upper arm circumference (MUAC), head circumference, weight-for-height (WHZ) |
| Bhaskaram et al. | 1986 | Confirmed cases (symptoms + serum antibody) | Kwashiorkor, marasmic kwashiorkor | Clinical signs and symptoms (details not specified) |
| Aaby & Coovadia | 1985 | Not specified | Protein-energy malnutrition (PEM) | Weight-for-height (WHZ), Height-for-age (HAZ), Weight-for-age (WAZ) |
| Oyedeji | 1984 | Not specified | Protein-energy malnutrition (PEM), Kwashiorkor, marasmus, marasmic kwashiorkor | Clinical signs and symptoms (weight loss, edema, skin, hair and psychological changes, emaciation with loss of muscle and fat, irritability) |
| Aaby et al. | 1984 | Not specified | Underweight | Weight-for-age (WAZ) |
| Lepage | 1983 | Not specified | Severe acute malnutrition | Harvard standards: Weight-for-height (WHZ), Height-for-age (HAZ) |
| Koster et al. | 1981 | Clinical signs and symptoms (rash) and serologic testing in one village | Protein-energy malnutrition (PEM) | Harvard standards: Weight-for-height (WHZ) or weight changes over a 2- months interval |
| Orren et al. | 1981 | Not specified | Not specified, marasmus | Boston standards: Weight-for-age (WAZ) |
| Whittle et al. | 1980 | Severe measles as defined by unusually severe inflammatory change in a tissue known to be infected with measles (ex.: extensive epidermal desquamation, severe keratitis, conjunctivitis, etc) | Kwashiorkor, marasmic kwashiorkor | Not specified |
| Whittle et al. | 1979 | Severe measles as defined by unusually severe inflammatory change in a tissue known to be infected with measles (ex.: extensive epidermal desquamation, severe keratitis, conjunctivitis, etc) | Kwashiorkor, marasmic kwashiorkor | Not specified |
| Dossetor et al. | 1977 | Clinical signs ands symptoms (rash) for sample selection; antigens testing for study | Not specified | Harvard standards: Weight-for-age (WAZ), Height-for-age (HAZ), serum albumin concentration |
| Dossetor & Whittle | 1975 | Clinical signs and symptoms (rash) | Underweight | Harvard standards: Weight-for-age (WAZ) |
| Axton | 1975 | Clinical signs and symptoms (rash) | Kwashiorkor | Not specified |
| Whittle et al. | 1973 | Severe measles as defined by unusually severe inflammatory change in a tissue known to be infected with measles (ex.: extensive epidermal desquamation, severe keratitis, conjunctivitis, etc) | Kwashiorkor, marasmic kwashiorkor | Not specified |
| Smythe et al. | 1971 | Not specified | Protein-calorie malnutrition (PCM) | Harvard standards: Weight-for-age (WAZ) |
| Morley | 1969 | Not specified | Not specified | Weight loss |
