## Supplementary material for "A scoping review on the associations and potential pathways between malnutrition and measles": S4 Table. Characteristics of vitamin A deficiency studies

| **S4 Table: Characteristics of vitamin A deficiency studies** | | | | |
| --- | --- | --- | --- | --- |
| **Authors** | **Date** | **Type of study** | **Setting** | **Indicator of vitamin A deficiency** |
| Hübschen et al. | 2022 | Review | Not specified | Not specified |
| Wang et al. | 2022 | Review | Not specified | Not specified |
| Strebel et al. | 2019 | Review | Not specified | Not specified |
| Asferaw et al. | 2017 | Descriptive | Community | Not specified |
| World Health Organization | 2017 | Review | Not specified | Not specified |
| Stevens et al. | 2015 | Analytical study: observational | Not specified | Serum retinol levels |
| Boushab et al. | 2015 | Descriptive study | Hospital | Serum retinol levels |
| Sachdeva et al. | 2011 | Analytical study: observational | Community | Clinical signs and symptoms: Bitot’s spot, corneal xerosis and ulceration / keratomalacia, corneal scar |
| Phillips et al. | 2004 | Analytical study: observational | Community | Serum retinol levels |
| Rosales | 2002 | Analytical study: experimental (RCT) | Hospital | Molar ratio of retinol-binding protein to transthyretin (RBP/TTR) |
| Khandait et al. | 2000 | Analytical study: observational | Community | Conjunctival impression cytology (enlarged epithelial cells, goblet cells and mucin spots absent) |
| Semba | 1994 | Review | Not specified | Serum retinol levels |
| Hussey & Klein | 1992 | Review | Hospital | Serum retinol levels |
| Caballero & Rice | 1992 | Review | Community | Serum retinol levels |
| De Sole et al. | 1987 | Analytical study: observational | Community | Clinical signs and symptoms: Bitot’s spot, corneal xerosis and ulceration / keratomalacia, corneal scar |
