## Supplementary material for "A scoping review on the associations and potential pathways between malnutrition and measles": S5 Table. Characteristics of vitamin A supplementation studies

| **S5 Table: Characteristics of vitamin A supplementation studies** | | | | | |
| --- | --- | --- | --- | --- | --- |
| **Authors** | **Date** | **Type of study** | **Setting** | **Type of vitamin A supplementation** | **Vitamin A dosage** |
| Hübschen et al. | 2022 | Review | Not specified | Therapeutic | Not specified |
| Lo Vecchio et al. | 2021 | Analytical study: observational | Hospital | Therapeutic | 2 doses of oil-based vitamin A; dosage based on child's age |
| Lowang et al. | 2021 | Analytical study: observational | Community | Preventative | Not specified |
| Domai et al. | 2021 | Analytical study: observational | Hospital | Therapeutic | Not specified |
| Hester et al. | 2020 | Analytical study: observational | Hospital | Therapeutic | 10,000 (62%) or 50,000 (13%) unit doses. No patients met current dosing recommendations for their age. |
| Strebel & Orenstein | 2019 | Review | Not specified | Therapeutic | Not specified |
| Imdad et al. | 2017 | Meta-Analysis | Community | Preventative | 10 000 IU to 200 000 IU; doses vary per study included in review (table 1) |
| Mafigiri et al. | 2017 | Analytical study: observational | Community | Therapeutic | Not specified (Y/N self-reporting survey) |
| World Health Organization | 2017 | Review | Not specified | Therapeutic | WHO guidelines |
| Bello et al. | 2016 | Systematic review | Hospital, Community | Therapeutic | Standard WHO dosage (54.5 mg for children < 12 months, 109 mg for children > 12 months) at admission and on days two, eight and week six in one study; single dose 200,000 IU in another study + 40 µG/mL vitamin E |
| Stevens et al. | 2015 | Analytical study: observational | Not specified | Preventative | Not specified |
| Mayo-Wilson et al. | 2011 | Meta-Analysis | Not specified | Preventative | 10 000 IU to 200 000 IU; doses vary per study included in review (table 1) |
| Mishra et al. | 2008 | Analytical study: observational | Community | Preventative | Not specified |
| Huiming et al. | 2005 | Meta-Analysis | Hospital, Community | Therapeutic | Doses vary per study included in the review |
| D'Souza RM & R D'Souza | 2002 | Meta-Analysis | Hospital, Community | Therapeutic | Doses vary per study included in the review (200,000 IU in a single dose or given on 2 consecutive days) |
| Rosales | 2002 | Analytical study: experimental (RCT) | Hospital | Therapeutic | 210 umol retinol as retinyl esters |
| Mahalanabis & Bhan | 2001 | Review | Not specified | Therapeutic | Doses vary per study included in the review (60 000 ug RE to 108 000 RE, cod liver oil) |
| Sommer | 1997 | Meta-Analysis | Hospital, Community | Preventative | Moderate to 200 000 IU vitamin A on 2 successive days (no specific amount for the moderate dose), or vitamin A fortified monosodium glutamate |
| Dollimore et al. | 1997 | Analytical study: experimental (RCT) | Community | Therapeutic | 100,000 IU of retinol equivalent for children aged 6-11 months or 200,000 IU for older children |
| Semba | 1994 | Review | Not specified | Therapeutic and preventative | Doses vary per review (200 000 IU vitamin A or 60 mg retinol equivalents, cod liver oil) |
| Madhulika et al. | 1994 | Analytical study: observational | Hospital | Therapeutic | 200 000 IU vitamin A on 2 successive days |
| Fawzi et al. | 1993 | Meta-Analysis | Hospital | Therapeutic and preventative | Doses vary per study included in the review (200 000 IU to 400 000 IU), divided in 2-3 doses, 300 Carr and Price units vitamin A and 2000 IU vitamin D |
| Hussey & Klein | 1992 | Review | Hospital | Therapeutic | 200,000 IU vitamin A given on 2 successive days, or 20,000 IU daily for 1-3 weeks |
| Barclay et al. | 1987 | Analytical study: experimental (RCT) | Hospital | Therapeutic | 200,000 IU vitamin A given on 2 successive days |
