## Supplementary material for "A scoping review on the associations and potential pathways between malnutrition and measles": S6 Table. Summary of methodological limitations in the existing evidence base

| **S6 Table: Summary of methodological limitations in the existing evidence base** | |
| --- | --- |
| Variation in or lack of definition for malnourished cases and/or measles cases  Variation in vitamin A supplementation dosage regimens (preventative and therapeutic) | Difficult to estimate a cumulative effect size for associations between the nutritional status and measles, and at higher risk of information biases (i.e. classification errors) |
| Observational designs were used in all studies reviewed that assessed the association between undernutrition and measles | Observational studies can be more vulnerable to biases and confounding factors, which can weaken the strength of inferences that we can draw from them |
| Use of proxy reports to identify malnourished and/or measles cases | Proxy reports are at higher risk of information biases (i.e. misclassifying a case) |
| Use of serum retinol to assess vitamin A status | Serum retinol can overestimate vitamin A deficiency prevalence in the presence of inflammation, which could bias the association observed to be stronger than it is |
| Lack of reporting, verification of potential effect modifiers and control of potential confounding variables in several studies (e.g. measles vaccination status, maternal nutrition, postnatal diet, breastfeeding practices, micronutrient deficiency status) | Analyses that do not adjust for the effect of potential confounding variables can bias study results and the measure of association estimated |
| Wide confidence intervals around the measure of association calculated in some studies | Wide confidence intervals indicate a higher degree of imprecision around the measure of association found |
